# Extrapolation of Infection Data for the CoVid-19 Virus in 21 Countries and States and Estimate of the Efficiency of Lock Down

**DOI:** 10.1101/2020.06.17.20134254

**Authors:** Walter Langel

## Abstract

Predictions about the further development of the Corona pandemic are of great public interest but many approaches demand a large number of country specific parameters and are not easily transferable. A special interest of simulations on the pandemic is to trace the effect of politics for reducing the virus spread, since these measures have had an enormous impact on economy and daily life.

Here a simple yet powerful algorithm is introduced for fitting the infection numbers by simple analytic functions. This way, the increase of the case numbers in periods with different regulations can be distinguished, and by extrapolating the fit functions, a forecast for the maximum numbers and time scales are possible. The effect of the restraints such as lock down are demonstrated by comparing the resulting infection history with the likely unconstrained virus spread, and it is shown that a delay of 1-4 weeks before imposing measures aiming at social distancing could have led to a complete infection of the respective populations.

The approach is simply transferable to many different states. Here data from six E.U. countries, the UK, Russia, two Asian countries, the USA and ten states inside the USA with significant case numbers are analyzed, and striking qualitative similarities are found.

Keywords: Covid-19, forecast, analytic fit, France, Germany, Italy, Spain South Korea, New York, Washington, Florida, Michigan, Poland, Sweden, USA, Pennsylvania, China, Russia, UK, California, Illinois, Indiana, Maryland, North Carolina.

## 1 Introduction

### 1.1 Issue

At the beginning in January, February, and beginning of March 2020, the total infection numbers showed a dramatic and grew nearly exponentially worldwide, and it seemed possible that finally a significant part of the population will be infected. By attaining herd immunity this way the virus should finally disappear.

It turned out that the mortality due to the virus was rather high. In the worst case, a death rate of e.g. 2% during infection of 2/3 of the population would result e.g. in Germany in 1.1 millions of victims before herd immunity was reached. At the same time, many health systems were overcharged and hospitals could no more provide adequate treatment to severely affected persons. As a consequence, herd immunity by rapid spreading of virus infection was no acceptable option, and many countries shutdown public life, schools and universities, shops and factories mostly in March. These measures extended for some weeks and slowed down infection rates, but caused enormous damage to economy and great social problems.

In the next stage pressure grew in the societies to release constraints step by step from May on, but parallel to that concern grows that this might lead to a second wave of infection spreading and that the pandemic might last for a long time and influence our daily life for years.

The details of shutdown and gradual release were subject to political decisions and to the specific situations in each state, and different ways have been followed in countries all over the world. It is thus of great interest to trace the virus spreading as a result of these measures quantitatively and inside a simple model, which makes data from different countries easily comparable. Obviously, it is also of great interest to have some forecast on the extension of the corona crisis.

### 1.2 Calculation methods

Beyond comparing data with an exponential increase, there are three approaches for understanding and predicting the development of case numbers:

Compartment models describe the kinetics of infection spreading by dividing the manifold of people into different groups of non-infected (N), susceptible (S), exposed (E), carrier (C), infected (I), recovered and dead (D). The definition of the compartments is somewhat arbitrary, and different models are around such as SIR (1) (2), SIRD (3), SECIR (4). Even small models need a fairly large number of parameters (4). It is demanding to define these parameters with sufficient precisions, and sophisticated statistical approaches are necessary (2) (1). Compartment models were applied to follow daily infection rates 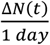. By several reasons, these data are strongly fluctuating, and compartment models may have to introduce additional assumptions to fit this fluctuation (1).

Sophisticated statistical models are used in epidemiology for evaluating the reproduction rate (5), but a recent application to data from European states showed that the data fluctuation also affects the model predictions (6).

Neuronal networks are a method of fitting data very precisely [ZHU], but the problem of this approach is that the number of parameters may be high. These parameters are hidden in the weights in the network, their values usually not having a well-defined physical meaning. It may thus be difficult to fit data from different countries and compare the parameters, and also to distinguish between noise and significant data. The authors of (7) conclude that this approach does not show advantages over analytic curve fitting.

A fit of the infection data by analytic function is tempting by three reasons

i. An analytical function can be chosen, which depends only on few parameters with a physical meaning. The fits provided here aim at determining two values, the predicted number of total infections *N*(*t* → ∞) at the end of the pandemic, when the infection rate has come down again, and the time scale, when this will happen.
ii. A fit of the infection data shall enable to distinguish between periods with different regulations such as before and during lock down, and the later release of shutdowns and social distancing. This is very difficult with fits trying to follow 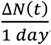, which are subject to enormous noise, especially weekly fluctuations (light blue in **Figure 1**), and this hides the effect of political measures in fits extending only over short times. Any interpretation of 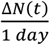 or even its change from day to day, which is the second derivative of *N*(*t*), is very risky. Mathematically, the total number of confirmed cases, *N*(*t*), is in turn the integral of 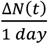 and thus behaves much smoother. It is, however, nearly impossible to distinguish between different exponential increase functions and the deviations from them by plotting *N*(*t*) on a linear y-scale. Analytical functions are not restrained to fitting the infection rate itself. Here the decadic logarithm *lg*(*N*(*t*)) of the total infection numbers is fitted by a smooth analytical function of time (dashed lines in Figure 1). Exponential behavior and deviation from that are easily interpreted. From the fit of *lg*(*N*(*t*)), calculated curves for *N*(*t*) and of 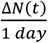 are obtained, which can be directly interpreted rather than the original zigzag data for the infection rate.
iii. Due to the transparent use of a small number of parameters, analytical functions are easily transferable between data sets from different countries and entities, showing convergence as well as differences. In this work the development in 21 countries and states is compared and a systematic classification is presented. The choice comprises two Asian countries, which very quickly extinguishing the virus spreading, six countries in the E.U., which were considered to be in similar conditions before the crisis broke out, but handled it quite differently, and Russia, the U.K. and the USA. In (8), ten states inside the USA are mentioned, which have high numbers of cases and thus could provide statistically significant data. According to (9), the data for states inside the USA indeed strongly diverge. Thus, data from the USA were not only treated as a whole, but also the numbers for ten states were analyzed separately.

**Figure 1:**
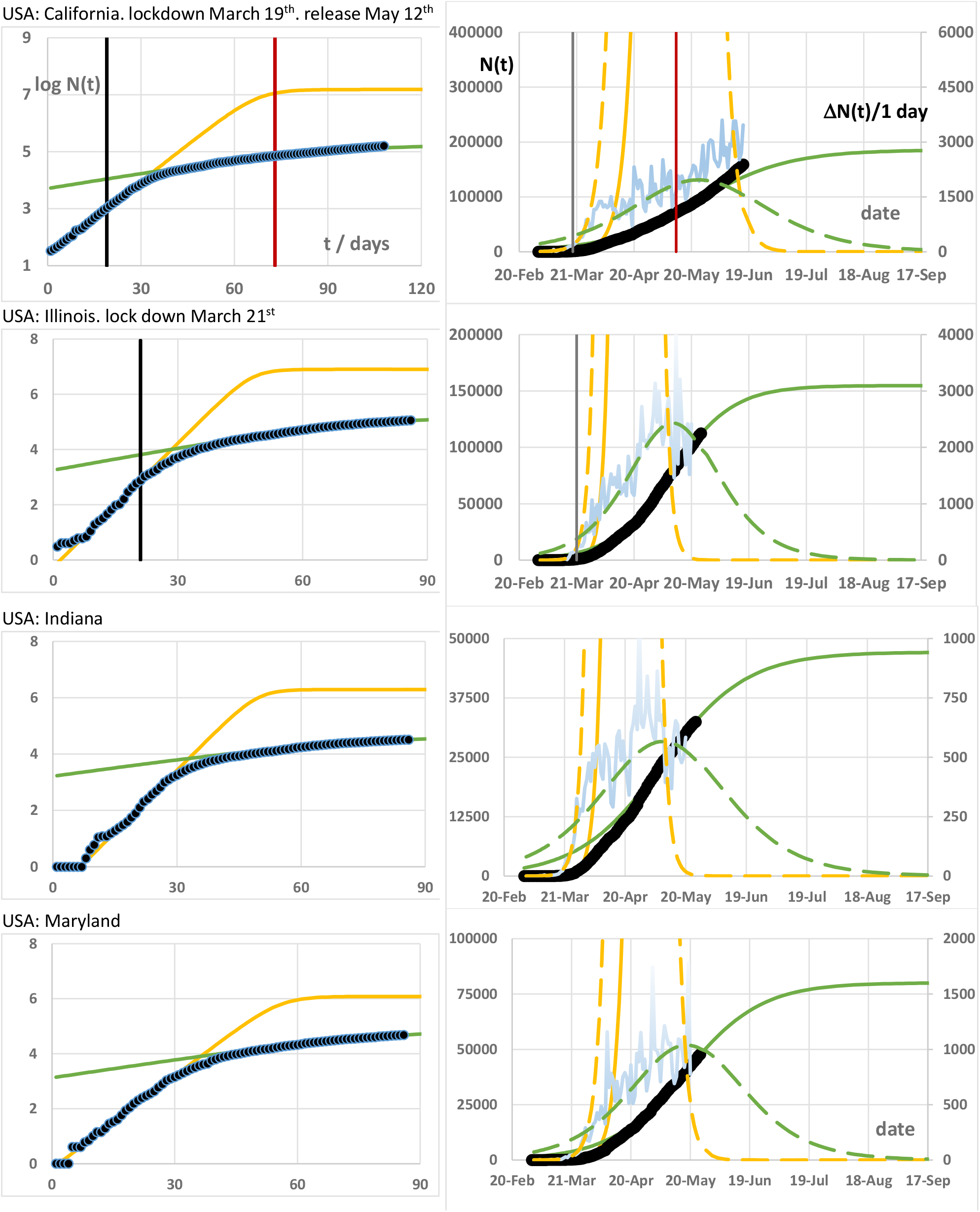
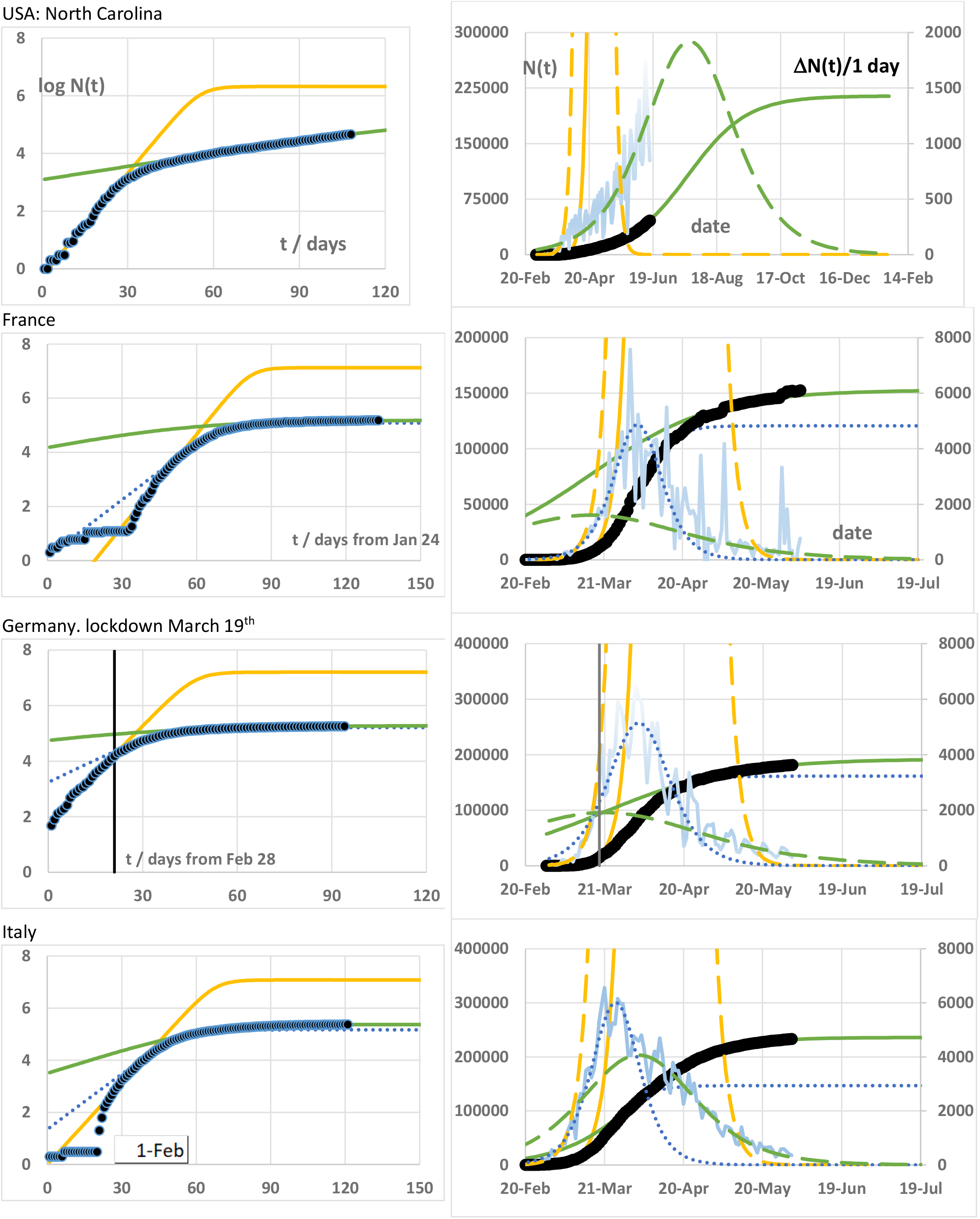
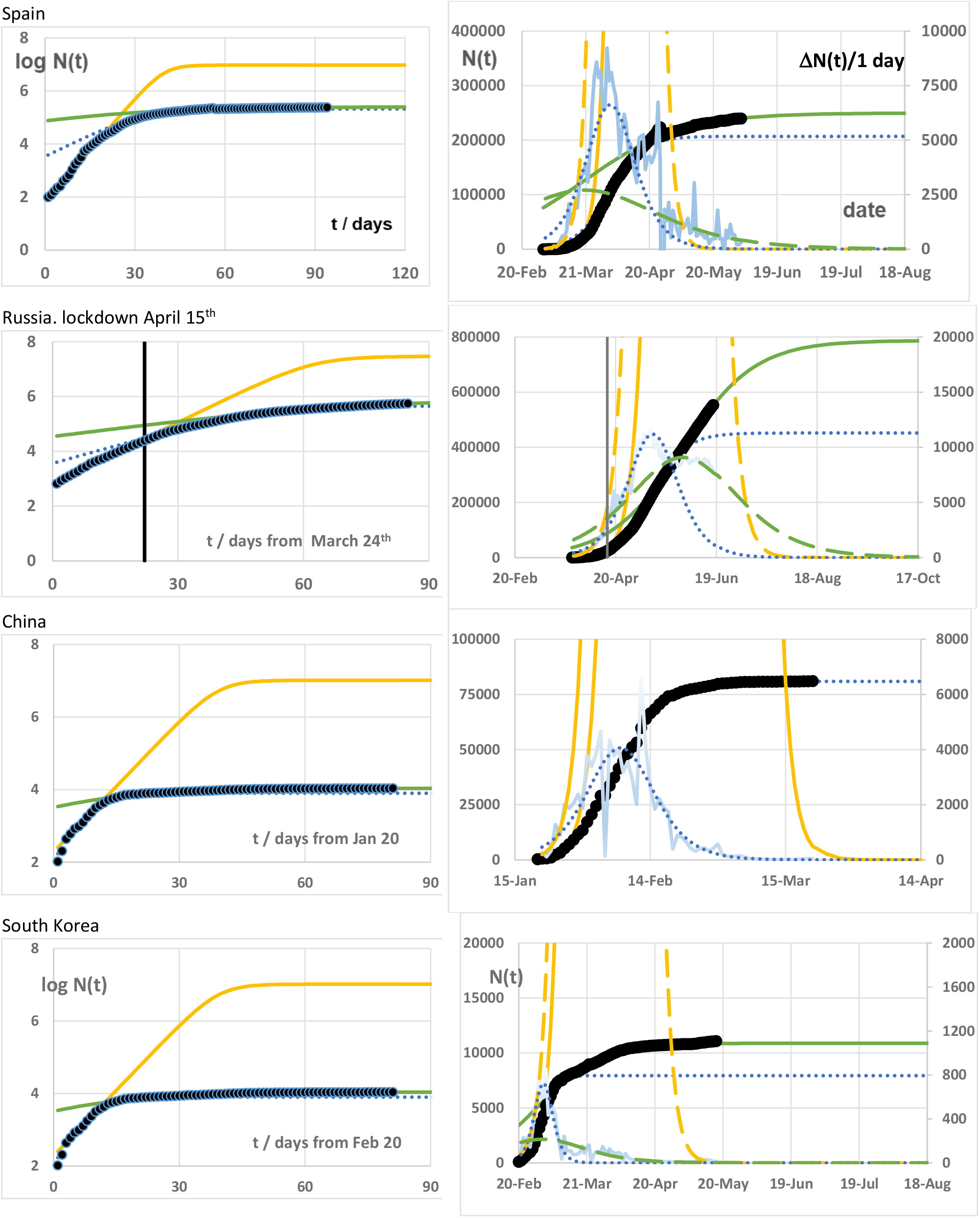
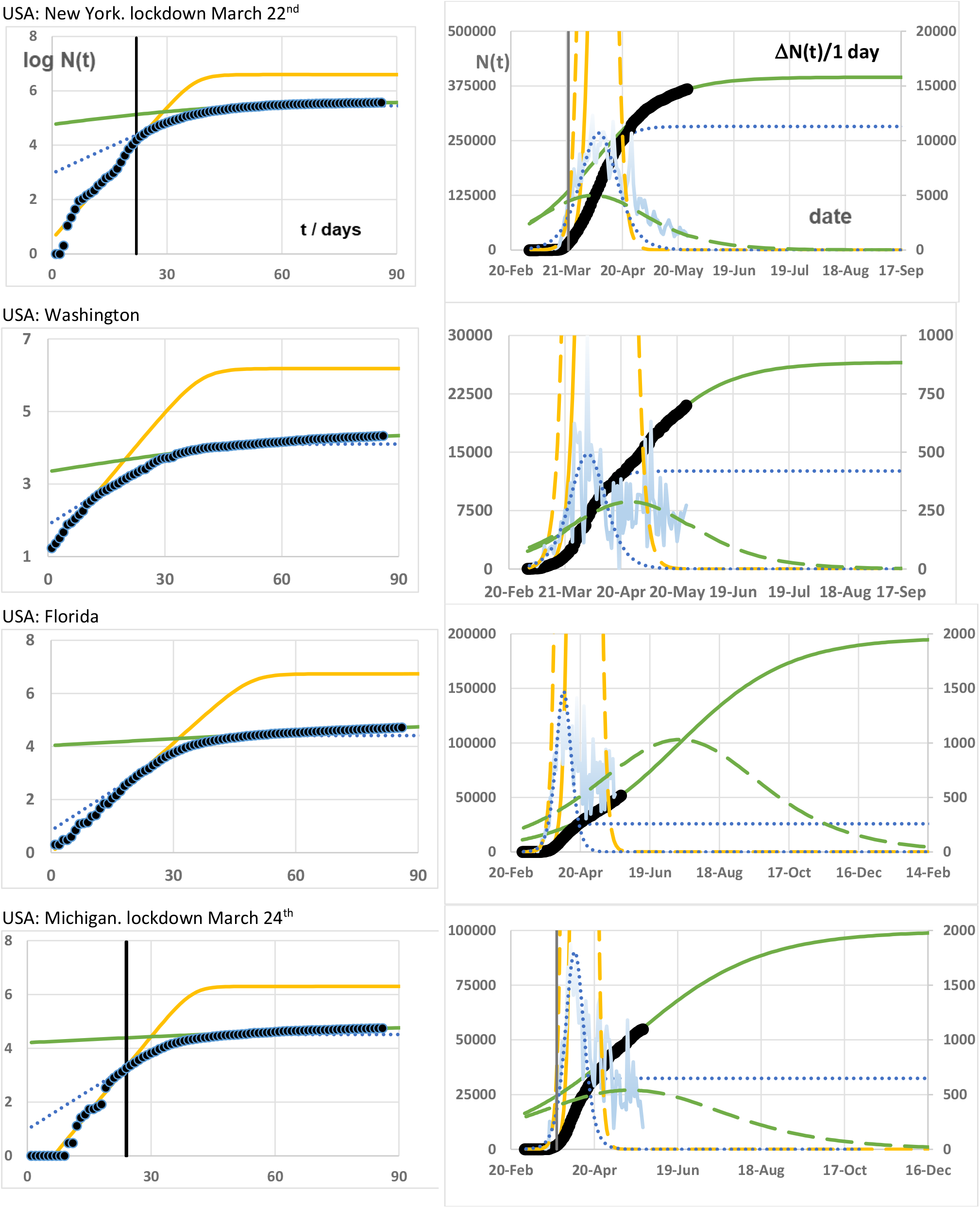
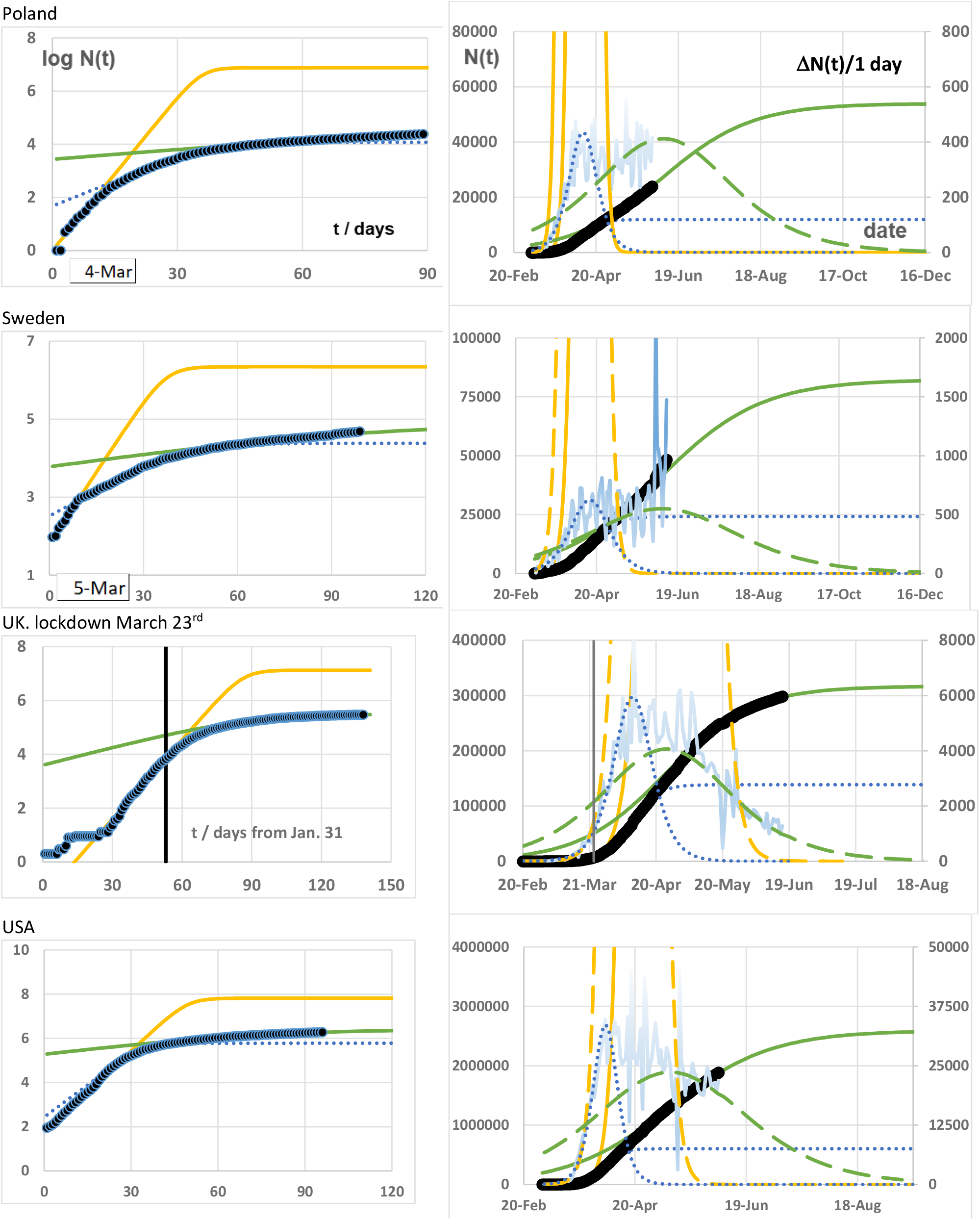
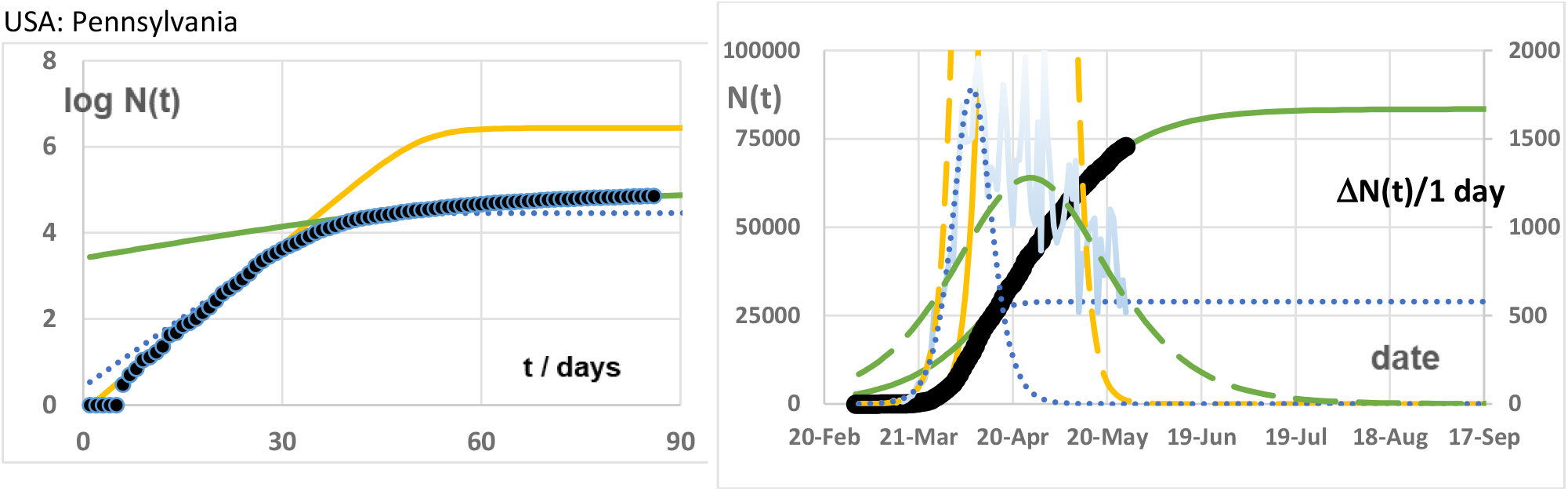
Data and fit results for the total confirmed case numbers in eleven countries and ten states of the USA. **Left column**: log-linear plots. X: Number of days since beginning of case report. Day 1 is March 1^st^ unless otherwise specified. Y: Decadic logarithm of total number of confirmed cases. (4 ⟺ 10,000, 6 ⟺ 1,000,000). **Right column**: X: date, Y: N(t). **Black full circles**: observed total case numbers. **Light blue lines**: daily infection rates. Orange. dotted blue and green lines are fits of data from different time windows (see text). Full and dashed lines refer to total numbers and rates. respectively.

The following results are used here for characterizing the virus spread:

- The maximum number of infected cases *N*(*t* → ∞), which is attained asymptotically by the fit function, characterizes the efficiency of the lock down.
- For comparison between different countries, the data for *N*(*t* → ∞) are also normalized inhabitants of the respective country/state yielding the “cumulative Incidence”. The term “incidence” is used here for describing normalization to 100,000 inhabitants, and “cumulative” means summed up till the end.
- The day *t*, at which the infection rate has its maximum value, 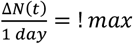 characterizes the time shift in different countries.

The length of the pandemic is characterized by two numbers:

- The fit parameter Δ*t*_10%_ gives the time in days until 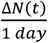 has dropped from its maximum value to 10% of it.
- Another instructive result is the date, at which this infection rate per day has dropped so far that the incidence per day is smaller than one independently of the previous maximum value. For Germany this corresponds e.g. to 830 new cases per day in 83 mill. of inhabitants, which means that the pandemic is not yet over, but controllable.
- A new time parameter Δ*t*_*hi*_ is introduced here with the following meaning: The lock down (fortunately) stopped the exponential increase before the full populations in the respective states were infected, and the case numbers then developed on a much smaller level. A delay of the lock down would have led to higher infection numbers and finally the whole population would have been infected, i.e. lock down would have had no more effect. Here, the time is estimated between actual lock down and the date, at which any measure would have lost its effect. It is seen that this time was only a few weeks long, and this figure demonstrates the importance of quick action for stopping the unrestricted spread of the virus.

At least two approaches have been presented so far. A power law approach has been presented in (10), but it is not clear how if the different sections with different functions reflect different periods of political measures for reducing virus spreading, or if the different analytical functions are just needed to describe one saturation function. In this case it may be difficult to fit consecutive saturation regimes this way.

In March 2020, at a time, when the lengths of shut down and pandemic was not yet provided elsewhere, a method was presented (11), which approximated the natural logarithm of the total case numbers by a logistic function. Very good fits were obtained, but as the number of data evolved, it turned out that the method did not reproduce obvious breaks between different periods sufficiently well. The present paper thus is based on the genuine logistic function (12) with again only three parameters. Even though the earlier predictions were based on much smaller data sets and used a different fit function, main conclusions are still consistent with the present calculation (**Table 1**). After reevaluating Δt_10%_ for the old fit, it turned out that the time scale was already fairly precise. The final number of total cases came out well in Germany, but was overestimated in Italy. Still it was already obvious from these fits that the maximum case number were orders of magnitude lower than the total population.

**Table 1:** Comparison of early fit (11) to the present data for the total case number and decrease of the rate to 10%:

|  | Germany |  | Italy |  |
| --- | --- | --- | --- | --- |
| Prediction using data until | $N(t \rightarrow \infty)$ | Date of 10% | $N(t \rightarrow \infty)$ | Date of 10% |
| March 24<sup>th</sup> | 221,000 | May 9<sup>th</sup> | 413,000 | May 28<sup>th</sup> |
| June 11<sup>th</sup> (s. below) | 192,000 | May 24<sup>th</sup> | 236,000 | May 20<sup>th</sup> |

## 2 Method

For eleven countries, the total case numbers were taken from (13), (14), and the infection rates were calculated as differences between total numbers of two following days. For the states of the USA, the infection rates per day were taken from (15), and the total numbers were obtained by summing up till the respective date.

A well-known formula for virus spread in a community provides the logistic function (12):

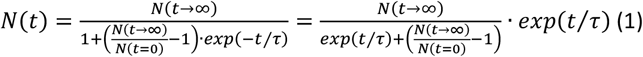

This formula only contains three parameters, the starting value *N*(*t* = 0) at *t* = 0, the final value *N*(*t* → ∞), which is attained at long times, and a time constant *τ*, which describes the transition from the exponential increase to saturation. This time parameter has a simple meaning, since Δ*t*_10%_ = 3.7 · *τ* is the time, in which the infection rate in a single fit decreases from maximum to 10% of the maximum value. -The function correctly reproduces the limiting asymptotic cases:

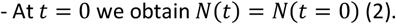

This starting point cannot be taken directly from some scattering initial data but is used as fit parameter. Its actual value depends on the time *t* = 0 when the data set starts.

For short times, while

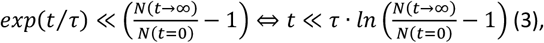

the formula reproduces a nearly exponential increase:

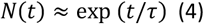

and a time for increasing by a factor of two given as

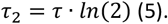

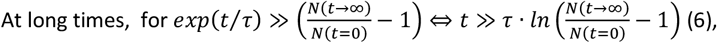

the function approaches asymptotically a maximum value, *N*(*t*) ≈ *N*(*t* → ∞).

This saturation behavior is generated by the exponential function in the first denominator in eq. (1) approaching zero for long times *t*. The length of the transition range from initial exponential increase to final saturation is described by the third parameter *τ*, which is characteristic for the rise time of the number of infections during initial exponential increase and also the saturation behavior.

Usually formula (1) describes the infection of a whole group with *N*(*t* → ∞) being equal to the total number of susceptible persons, and the members of the group finally obtaining herd immunity. Here, such a high limiting value applies only to the very first stage of virus spreading without any intervention. As the effect of measures is to be traced, it is not sensible to fit the whole time range of the pandemic by a single function. On the other hand, fitting too small intervals would introduce large errors due to the noise of the data. Instead, three time windows are defined for nearly homogeneous periods and fitted separately. The intervals are different for the different countries and do not necessarily coincide with dates of political measures. The dates may roughly correspond to initial free spread of the virus, short time effect of strict lock down and longtime behavior of the infection numbers. For describing the effect of lock downs, eq. (1) is used in slightly different way than originally. *N*(*t* → ∞) is now a fit parameter having values of only a small fraction of the population, such as several ten or 100 thousand people.

For the fit of equation (1) to the data points, the standard solver in Microsoft Excel 2016 was applied to the logarithm of the number *N*(*t*) of confirmed infections as saved in steps of one day, and the least squares error with respect to equation (1) was minimized by varying the three parameters *N*(*t* = 0), *N*(*t* → ∞), and *τ*. The data were weighted by 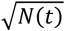 to account for poor statistics of small numbers.

## 3 Results

### 3.1 General features

There are important similarities between the fits for all countries and states considered here (**Figure 1**). In the log-linear plots, a sharp concave bending is seen somewhere between late March and early April. This effect is obvious without any fitting model, and one could essentially even draw straight lines with different slopes through the data points before and after this turning point. This bending is ascribed to the effects of any social distancing, which interrupted the originally exponential increase of infection numbers at around the same time in many countries. For some plots in **Figure 1**, (Germany, California, Michigan, New York, Russia, UK), vertical lines indicate the known dates of lock downs. In Germany, UK, Michigan and New York, the bend can be assigned to the respective lock down, since the observed total case numbers start to fall below the orange fit shortly afterwards. In California and Russia, the situation is less clear, since strict lock downs took place already on March 19^th^ (16) and 15^th^ (17), respectively.

The fitting periods were handled as follows:

1. Data during the time before intervention rose very quickly in all states. Orange lines were fitted to only a few points from the first days in this time interval, and an essentially linear increase is seen on the logarithmic scale. This means that the increase was still exponential at that time, and probably the whole population would have been infected in a short delay without precautions. In the range of exponential increase the slope m in the log-linear plot, the initial doubling time *τ*_2,*free*_ and the parameter *τ* of the fit function are related by

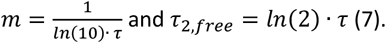 For *τ*_2,*free*_, small values between only two and six days were found (**Table 2**), and this has largely contributed to the fear, which has raised the appearance of CoVid-19. The final saturation value for the case number without intervention cannot be extracted reliably from this fit. Here an estimate of 20% of the respective population is used. This is significantly below 100% since there is probably a high number of unreported cases, and herd immunity is attained at an infection of significantly less than the full population (18). As the saturation value is not known with any precision, the horizontal parts of the orange lines are considered as guides to the eye. On the other hand, it is of little relevance to speculate about the saturation value of the infection numbers without retarding measures.
2. In all cases considered here, significant deviations from the initial exponential increases have been found already after a few days.

**Table 2:** Simulation results for ten countries including the USA and eleven states inside the USA. Data were extracted from the fits in **Figure 1** and arranged according to the fit type only, but disregarding political correlations and the scale of the infection in each country. The time unit is days. The columns contain: -sizes of populations *P* (20): Eventual uncertainties in these numbers are probably small as compared to errors in the fits and the resulting maximum numbers of cases. -The initial doubling time *τ*_2,*free*_ during uncontrolled virus spreading is related to the initial slope of the orange fit. -*N*(*t* → ∞), the extrapolated maximum case number (cf. full green line in **Figure 1**) -Δt_*hi*_, the efficient time, by which the virus spreading was slowed down before large scale virus spreading would have taken place with final herd immunity, but with an uncontrollable death number. -The fit types are denoted F1, F2 and F3 according to the importance of the fitting by the short time (dotted blue) or long time (full and dashed green) functions (s. text and **Figure 1**). -*τ*_*bl*_ and *τ*_*gn*_ are the time constants for the fits by blue and green lines, respectively. The importance of these constants is readily seen by inspecting the respective results for the infection rates in the linear plots. -New cases per day: Figures in the next four columns reflect the importance and duration of the pandemic in each country: (1) Maximum new infections per day, (2) respective date, (3) date, when the rate has decreased to 10% of its maximum, and (4) time between these two dates. -Extrapolated cumulative incidence. -Incidence per day <1: An estimate for the duration of the pandemic is given by evaluating the date, when the incidence has dropped below 1 case per day.

|  |  |  |  |  |  |  |  | New cases per day |  |  |  |  |  |
| --- | --- | --- | --- | --- | --- | --- | --- | --- | --- | --- | --- | --- | --- |
| | $P$ | | | | fit type | | | Maximum | | <10% of peak | | Extrapolated Cumulative incidence | Incidence per day <1 |
| | | $\tau_{2,free}$ | $N(t \rightarrow \infty)$ | $\Delta t_{hi}$ | | $\tau(bl)$ | $\tau(gn)$ | at | value | after | $\Delta t_{10\%}$ | | after |
| California | 39,300,000 | 3.5 | 186,000 | 19 | F1 |  | 24 | 24-May | 1964 | 21-Aug | 89 | 472 | 3-Aug |
| Illinois | 12,670,000 | 2.0 | 155,000 | 8 | F1 |  | 16 | 10-May | 2432 | 9-Jul | 60 | 1222 | 21-Jul |
| Indiana | 6,732,000 | 2.1 | 47,000 | 10 | F1 |  | 21 | 9-May | 567 | 27-Jul | 79 | 700 | 21-Jul |
| Maryland | 6,046,000 | 2.7 | 80,000 | 11 | F1 |  | 19 | 18-May | 1038 | 30-Jul | 73 | 1324 | 11-Aug |
| North Carolina | 10,490,000 | 2.5 | 214,000 | 8 | F1 |  | 28 | 14-Jun | 1238 | 16-Nov | 155 | 2042 | 22-Nov |
| France | 66,990,000 | 2.6 | 153,000 | 17 | F2 | 6.2 | 24 | 3-Apr | 4875 | 13-May | 40 | 228 | 3-May |
| Germany | 83.100.000 | 2.6 | 192.000 | 17 | F2 | 7.8 | 25 | 3-Apr | 5143 | 24-May | 51 | 231 | 10-May |
| Italy | 60.360.000 | 2.9 | 236.000 | 16 | F2 | 6.1 | 14 | 25-Mar | 6008 | 20-May | 56 | 391 | 20-May |
| Spain | 46.940.000 | 2.3 | 250.000 | 12 | F2 | 7.8 | 23 | 1-Apr | 6613 | 21-May | 50 | 532 | 30-May |
| Russia | 146.570.133 | 4.1 | 787.000 | 21 | F2 | 10.1 | 22 | 12-May | 11199 | 16-Aug | 96 | 537 | 7-Aug |
| China | 1.393.000.000 | 1.6 | 81.000 | 19 | F2 | 5.0 | 5 | 7-Feb | 4060 | 26-Feb | 19 | 6 |  |
| South Korea | 51.640.000 | 2.5 | 11.000 | 25 | F2 | 2.7 | 13 | 2-Mar | 726 | 31-Mar | 29 | 21 |  |
| New York | 19.750.000 | 1.9 | 395.000 | 6 | F2 | 6.6 | 20 | 7-Apr | 10696 | 3-Jun | 57 | 2000 | 6-Jul |
| Washington | 7.615.000 | 2.3 | 27.000 | 14 | F2 | 6.4 | 23 | 2-Apr | 493 | 6-Jul | 95 | 349 | 27-Jun |
| Florida | 20.980.000 | 2.2 | 197.000 | 10 | F2 | 4.4 | 48 | 6-Apr | 1472 | 18-Dec | 256 | 938 | 30-Nov |
| Michigan | 9.987.000 | 1.6 | 100.000 | 7 | F2 | 4.5 | 46 | 3-Apr | 1806 | 26-Aug | 145 | 999 | 1-Oct |
| Poland | 37.970.000 | 1.6 | 54.000 | 11 | F3 | 6.9 | 33 | 11-Apr | 434 | 4-Oct | 176 | 142 | 30-Jun |
| Sweden | 10.300.000 | 2.5 | 82.000 | 12 | F3 | 9.7 | 37 | 15-Apr | 624 | 6-Oct | 174 | 798 | 27-Sep |
| UK | 66.650.000 | 3.1 | 317.000 | 17 | F3 | 5.8 | 19 | 10-Apr | 5945 | 4-Jul | 85 | 475 | 28-Jun |
| USA | 328.200.000 | 2.5 | 2591.000 | 11 | F3 | 4.5 | 27 | 4-Apr | 33636 | 9-Aug | 127 | 789 | 9-Aug |
| Pennsylvania | 12.800.000 | 2.3 | 84.000 | 11 | F3 | 4.0 | 16 | 7-Apr | 1789 | 19-Jun | 73 | 654 | 28-Jun |

The full green lines (**Figure 1**) describe the increase of total cases after the lock down and release since beginning May up to June 6^th^. At the first glance they look as straight lines with a much smaller slope than for the orange lines in their exponential regime. This would still correspond to an exponential increase, which is just slower than before lock down.

Closer inspection shows that the green lines are slightly concave and thus increase slower than exponentially. Here, from the deviation of the case numbers from a strictly exponential increase during lock down and especially for the present release period, well defined fits are obtained. They permit to quantify the reactions to limitations such as social distancing. The fits yield predictions of end of the infection period and of the total case numbers *N*(*t* → ∞) (**Table 2**). These fits with a single broad logistic functions (green in **Figure 1**) with maxima between end of April and end of July spreading over a somewhat longer time are denoted as F1. The time Δt_10%_ between maximum of the infection rate and its decrease to 10% of the maximum is as long as 60-100 days.

The vertical positions of the green fits directly depend on the dates, when interventions took place. Lock down at earlier times resulted in a lower lying line and a smaller maximum number of total cases, whereas imposing the lock down later led to an increase of *N*(*t* → ∞). After waiting another time Δ*t*_*hi*_, the green line would have converged to the horizontal orange line meaning full infection. As was explained above, the herd immunity is estimated to be reached, when the total number of reported cases attains 20% of the population *P* in the respective country. We thus obtain

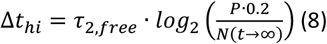

The doubling times *τ*_2,*free*_ of about 1.5-4 days for the exponential increases before lock down were taken from the slopes of the full orange lines in **Figure 1**. The estimate for Δt_*hi*_ was made as follows e.g. for France (cf. **Table 2**): Herd immunity is reached at *N*_*hi*_ = 0.2 · 66.990.000 = 13.397.500 reported cases, but in fact the total infection number will rise to *N*(*t* → ∞) = 153.00 cases. We thus obtain 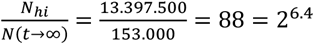. By waiting 6.4 doubling times of *τ*_2,*free*_ = 2.6 *days*, i.e. Δt_*hi*_ = 6.4 · 2.6 *days* = 17 *days* longer until imposing a lock down, one would have attained a number of reported cases equal to the estimated full infection with 13.4 mill. reported cases, rather than 153,000 after lock down. A delay by only two and a half weeks would have had the result that the green curve had converged to the orange curve, i.e. the lock down would not have had any effect anymore. In all cases, Δ*t*_*hi*_ is only between one and less than four weeks (**Table 2**). This demonstrates, how important it had been to rapidly decide appropriate measures in the respective countries.

The eq. (1) was fitted to the logarithm of the total numbers, but additional information is obtained from the linear plots (**Figure 1**, right column). The full green lines fit major parts of the data points for *N*(*t*), mostly since beginning of May, and are equivalent to the dashed green line for Δ*N*(*t*)/1*day* for new infections, which match especially the infection rates in the time interval (light blue lines in **Figure 1**). Other than from the directly observed noisy data, one obtains a time scale from the fit, and sees clearly, when the infections rise to a maximum and decrease again.

In many plots, the intermediate data points between the initial free virus spread and the slower infection rates after gradual release are not satisfactorily fitted by the green lines. The data from many countries in this intermediate range in the log-linear plots are well fitted by a third set of parameters (dotted blue lines), which describes the curvature of data between steep initial growth and slower increase after the lock down. These dotted lines predict lower total case numbers and a shorter time scale than the green lines (**Table 2**). Fit types providing this additional short time function are referred to as F2 and F3. This function with time constants *τ*_*bl*_ (dotted blue) of 3-10 days may be assigned to strict lock down measures.

It is seen now that in all considered countries and states the infection rate is not yet zero. The measures have nowhere full extinguished the virus spread, and the total numbers of cases still increase, may be enhanced by relaxation of the strict lock down. In addition to the short time fit described above, the longtime behavior for F2 and F3 still has to be described by a contribution on an extended time scale with *τ*_*gn*_ around 10-40 days. Only for the data for China available so far, no sensible longtime fit could be calculated. This *τ*_*gn*_ is longer than *τ*_*bl*_ and of the same order of magnitude as the time constants for the only function in F1. Due to the superposition of the two short and long time functions, Δt_10%_ scatters strongly for F2 and F3 fits, and values of 19 to 256 days were found without obvious correlation to other parameters.

The difference between these types is, how much this long time fit (green lines in Figure 1) contributes to the total case number:

F2: For a number of countries including France, Germany and Spain (cf. **Table 2** and **Figure 1**), the dashed green line only adds a small number of new cases probably due to a careful release of the restrictions, and the full green line is only slightly higher than the predictions from the short time fit, meaning that the total number of cases only slightly increases. Probably the release has only slightly prolongated the infection period and raised the total number.

F3: In a series of other countries, the longtime fit yields a significant contribution to the total number of cases, as the infection rate remains high. Only in some cases (e.g. Poland) a clear distinction between the first and second period is made, in other examples (USA), a nearly constant infection rate is observed so far. An exception is Sweden, where the rate dramatically increases since June 1^st^, and the data would be consistent with a second exponential increase only.

The following parameters characterize the time scale (**Table 2**).

i. The date of maximum infection rate, which was already passed in many countries at the beginning of April, in China even in February.
ii. The date, at which the infection rate has dropped below 1 case per 100,000 per day. It is seen that the infections will at least in some states (Florida, North Carolina) persist till end 2020.

### 3.2 Results for individual countries and states

As this paper is mainly on demonstrating the value of a simple analytical fit method, no deeper discussion on special countries will be given, but only a few remarks are made.

#### 3.2.1 Asia

As is well known, the Asian countries China and South Korea have extremely low infection numbers as compared to their large populations, and the extrapolations result in small cumulated incidences of six and 21 cases, respectively. Both countries obviously have strictly reacted to the pandemic, and the fit types are F2 with a strong short time component. None of them ever reported incidences above one. In the case of China it has to be considered that localized strong eruptions are normalized to a very large population. South Korea obviously was very quick in reacting to the pandemic, which is demonstrated by the longest time found here for any country of Δt_*hi*_ = 25 *days* (**Table 2**).

#### 3.2.2 Europe

##### 3.2.2.1 European Union

Even though the EU countries were closely connected before the pandemic and reacted on a similar time scale, the development in Europe diverged. For the four countries France, Germany, Italy, and Spain, similar parameters were found (**Table 2**), and the fit type was F2, reflecting a strong impact of the lock down measures in these countries. In Italy the long time contribution to the case number is rather high, even though the country had at least as strict contact bans as the other three. The extrapolated cumulative incidence ranges from 228 to 532, i.e. is in Spain higher by a factor of two than in France. The incidence per day dropped below one in May, which makes the opening of these countries for tourism plausible.

The severe problems, Spain France and Italy experienced during the initial phase of the pandemic, were not due to high average cumulative incidences, but to localized very high infection numbers and initial problems in the local health systems.

In the two further EU countries included in this study, Poland and Sweden, the impact of the measures was less obvious, and the fit type F3 indicates an important long time contribution to the cumulative incidence. The time scales and absolute case numbers are quite similar in both countries. Δt_10%_ is as long 120 and 141 days, as compared to 40-60 days in the four EU countries mentioned above, and the infection rate will drop to 10% in October rather than in May. As Sweden is much smaller than Poland, its resulting extrapolated cumulative incidence (840) is very high, and very low in Poland (140), and the barrier of 1 new case per day will be reached much earlier in Poland than in Sweden (end July vs. October). It may be speculated if the liberal contact ban policy in Sweden and the early strict closing of the borders in Poland, respectively, have incited these diverging results.

Due to the long time scale, both countries are still at the beginning of the pandemic, and predictions have to be taken with care. Especially in Sweden, the high case numbers since June 1^st^ – 11^th^ are also compatible with a new uncontrolled exponential rise.

##### 3.2.2.2 Russia and UK

The number of total cases *N*(*t* → ∞) may reach 800,000 in Russia vs. more than 300,000 in the UK, but the extrapolated cumulative incidences then will be in the same range around 500 for both countries (**Table 2**). Both infections rates have passed the maxima and start to decrease, even if this is partially hidden by fluctuations, and both data sets have a short time contribution, being more pronounced for Russia (F2) than for the UK (F3), and strong long time contributions have to be expected. Apart of that the time histories are very different:

The total case numbers in the UK showed a clear reaction to the lock down starting March 23^rd^, which is still at least in part maintained (**Figure 1**). The maximum of new infections per day will decrease rather fast, and the incidence of 1 might be reached already beginning of June in the UK, but only in August in Russia.

The log-linear plot for Russia shows a significant curvature, suggesting that the number of confirmed infection cases was very early increasing significantly less than exponential. A continuously concave dependence of *log*(*N*(*t*)) over *t* is observed (**Figure 1**). This indicates the superposition of the effect of successive measures starting with a lock down after March 30^th^. The digital pass system for Moscow starting on April 15^th^ is indicated in the plot, suggesting that this measure was the most efficient. No effect of release is visible in the data plotted.

#### 3.2.3 USA

The total incidence for the whole USA is around 800, which is at the upper end of the European states. A closer inspection of ten states in the USA showed that the number for the whole country are averaged over states with similarly case numbers but very different time histories. Even the fit types are different, and all three defined types occur (**Table 2**).

The extrapolated cumulative incidences for ten states inside the USA with high case numbers are compared with the policy in the respective state (8) in **Table 3**. The three states, which “monitor vulnerable populations”, have a significantly lower number of cases to be expected than the others. Even Pennsylvania with its large cities still has a lower value than Indiana. A very rough estimate is that this monitoring reduces the case number in comparable countries by a factor of two to three. This would be consistent with the experience in Europe that residences of elder people are very severely affected by the virus.

**Table 3:** Criteria considered for reopening by ten American states with high infection rates and respective incidence (8) are juxtaposed with rounded fit results in the present work for the predicted total incidence *N*(*t* → ∞) at the end of the pandemic. Countries monitoring the vulnerable populations show significantly lower incidences than states. which do not. Even Pennsylvania with its large industrial sites. which might be more susceptible to high infection rates. is only comparable to Indiana with a more agricultural structure (21). Washington and California have rates comparable to EU countries. the other states of the USA probably have lower incidences.

| State | Control virus spread | Ensure hospital capacity | Contain new cases | Monitor vulnerable populations | Rounded Cumulative Incidence |
| --- | --- | --- | --- | --- | --- |
| California | x | x | x | x | 500 |
| Florida | x | x | x |  | 1000 |
| Illinois | x | x | x |  | 1000 |
| Indiana | x | x | x |  | 700 |
| Maryland |  | x | x |  | 1000 |
| Michigan | x | x | x |  | 1000 |
| New York | x | x | x |  | 2000 |
| North Carolina | x |  |  |  | 2000 |
| Pennsylvania | x |  | x | x | 700 |
| Washington | X | x | x | x | 500 |

Two extremely diverging cases are California and North Carolina with 470 and 2040 extrapolated total incidence, respectively.

Case numbers in California first showed the exponential increase (orange in **Figure 1**) as seen everywhere. Around March 19^th^ a lockdown became effective, and the case number was significantly lower than the initial exponential curve would have predicted. A single fit (F1) resulted in 186,000 extrapolated total cases and an incidence of 470, which is well in the range of many European countries. By May 12^th^ (16) the lockdown was released to some extent. As a striking result, the new cases per day started to increase again strongly on a nearly exponential line well beyond the original fit. On the other hand, data for North Carolina yielded a high number of infections from the beginning. The maximum of daily infections is only reached end of July, and one has to wait till beginning of November, until the infection rate is decreased to 10% of the peak value. North Carolina is described as a more agricultural state (19), and should not have to fight infections in large industrial centers as do New York, Illinois, California and Pennsylvania. The high number of infections in this state thus seems to be remarkable and may be related to the fact, that N. C. is the only state among the ten not containing new cases (8).

## 4 Conclusion

The intention of the present manuscript is to present a model, which permits to trace the Corona virus spread as a function of interventions and to quantify effects on the basis of empirical data and few significant fit parameters. Such models are of great relevance for giving a justification for imposing or releasing measures such as contact ban or lockdown, which have a huge worldwide impact on the economy and daily life. This work is complementary to extended studies using epidemiological models with a large overhead, which are often difficult to apply and to transfers. It has been shown that the approach followed here permits the straightforward comparison of data from different countries.

I want to make the following points:

1. Log-linear plots indicate very clearly the exponential rise of infection case numbers and deviation from it as a consequence of contact ban and lock down. The deviation may often be correlated with political dates
2. An analytical fit with few parameters is attained by using a logistic function, where the asymptotically reached final value is used as fit parameter and yields the maximum number of infected individuals. This function reproduces the typical saturation behavior of infections. The timescale of this saturation is determined by a single time constant.
3. The deviation from the initially exponential increase usually as effect of political measures quantified by a newly introduced parameter Δt_*hi*_, which reflects the time between intervention and reaching full infection and herd immunity. This time was often only 1-2 weeks in the present crisis.
4. The reaction to the interventions is fitted by a short time and a long time function. In many cases, the first reflects the immediate response to lockdowns, the second one a long time development, e.g. after release of contact bans. Fit type short maximum, late maximum, exp increase. The release leads to an increase of the infection number, but the effect is different in strength for different countries. In Germany and France, rather small longtime fits describe the increase, but in Sweden and California the reaction to the release is an uncontrolled exponential increase of the case number and infection rate. Such examples demonstrate that the applied analytical fit method permits a rapid comparison of different states and data sets.
5. In the most cases, the time dependence of case number and infection rate can be fitted by saturation functions. This fit is quantified by two time parameters, the decrease of the infection rate from its maximum to 10% of its value, and the time, when the normalized infection rate in a country reaches 1 per 100,000 per day.
6. This analysis permits a forecast for the final incidence and the duration of the pandemic by extrapolation of fit functions. Moreover, one rapidly notices effects such as the uncontrolled increase of case numbers by monitoring sudden deviations from the fit.
7. For ten states inside the USA, a correlation between the criteria for reopening and the calculated cumulative incidence was found. California, Pennsylvania and Washington, which monitor vulnerable populations, had lower predicted values. North Carolina does not contain new cases and has a very high calculated value.

## Data Availability

The full excel file containing data and fits will be made available on request. Data used were taken from public ressources.

